# Association of effective circulating blood volume with sublingual RBC velocity and microvessel pressure difference in anesthetized individuals: A clinical investigation and computational fluid dynamics modeling

**DOI:** 10.1101/2022.05.09.22274826

**Authors:** Athanasios Chalkias, Michalis Xenos

## Abstract

**Background:** Although changes in effective circulatory volume may affect microcirculatory red blood cell (RBC) velocity and oxygen extraction ratio, no systemic variable has been consistently associated with hemodynamic coherence. We therefore evaluated the association between mean circulatory filling pressure and microcirculatory perfusion and oxygenation.

**Methods:** This analysis included anesthetized individuals in steady-state physiology. We assessed the correlation of mean circulatory filling pressure analogue (Pmca) with sublingual microcirculation and RBC velocity using SDF+ imaging and a modified optical flow-based algorithm. We also reconstructed the 2D microvessels and applied Computational Fluid Dynamics (CFD) to evaluate the correlation of Pmca and RBC velocity with the obtained pressure and velocity fields in microvessels from CFD [pressure difference (Δp)].

**Results:** Twenty adults were included in the study, of whom 12 (60%) were men and 8 (40%) were women, with a median age of 39.5 years (IQR 35.5-44.5). Sublingual velocity distributions were similar and followed a log-normal distribution. A constant Pmca value of 14 mmHg was observed in all individuals with sublingual RBC velocity of 6-24 μm sec^-1^, while a Pmca <14 mmHg was observed in those with RBC velocity >24 μm s^-1^. When Pmca ranged between 11 mmHg and 15 mmHg, Δp fluctuated between 0.02 Pa and 0.1 Pa.

**Conclusions:** These data suggest that the intact regulatory mechanisms may maintain a physiological coupling between systemic hemodynamics and tissue perfusion and oxygenation when Pmca is 14 mmHg.

**HIGHLIGHTS:**

- Changes in effective circulatory volume may affect microcirculatory red blood cell (RBC) velocity and oxygen extraction ratio, but no systemic variable has been consistently associated with hemodynamic coherence
- We found that a mean circulatory filling pressure analogue (Pmca) value of 14 mmHg maintains sublingual RBC velocity at 6-24 μm s^-1^
- A Pmca <14 mmHg is observed in individuals with RBC velocity >24 μm s^-1^
- Microvessel pressure difference is maintained constant when Pmca ranges between 11 mmHg and 15 mmHg
- These findings imply that tissue oxygenation can be maintained in very low RBC velocities
- Therapeutic strategies and translational research beyond optimizing macrohemodynamics and microcirculatory flow are required, including normalizing red blood cell velocity

**BRIEF COMMENTARY:** *Background:* Hemodynamic coherence is not well-investigated and no systemic hemodynamic variable has been consistently correlated with microcirculatory perfusion and tissue oxygenation.

*Translational Significance:* The present study provides a novel method to monitor hemodynamic coherence and tissue perfusion, which can aid in the identification of novel hemodynamic phenotypes and enhance microcirculation-guided therapeutic strategies, optimizing local delivery of oxygen. Our findings also imply that (1) tissue oxygenation can be maintained in very low red blood cell velocities; and (2) therapeutic strategies and translational research beyond optimizing macrohemodynamics and microcirculatory flow are required, including normalizing red blood cell velocity.

## 1. INTRODUCTION

Physiological hemodynamic coherence is the condition in which the systemic hemodynamic variables are translated into effective microcirculatory perfusion and oxygen delivery to the parenchymal cells. This requires normal physiology and intact compensatory mechanisms to regulate oxygen transport to tissue. Although hemodynamic coherence was first described in 1850, only recently has it been studied in critically ill patients. Nevertheless, its characteristics in steady-states are not well-investigated and no systemic hemodynamic variable has been consistently correlated with microcirculatory perfusion and tissue oxygenation.

As the main role of the venous system is to serve as a capacitance to maintain filling of the heart and cardiac output (CO), assessment of mean circulatory filling pressure (Pmcf) is a basic parameter of functional hemodynamic monitoring. Mean circulatory filling pressure is the blood pressure throughout the vascular system at zero flow [1]. Based on a Guytonian model of the systemic circulation, an analogue of Pmcf (Pmca) can be derived that adequately follows intravascular volume status [2,3], its measurements are automatic, and can characterize the hemodynamic response to treatment modalities [4,5].

Decreases in effective circulatory volume may affect microvascular red blood cell (RBC) velocity resulting in insufficient oxygen extraction ratio (O_2_ER) [6,7]. On the other hand, fluid or vasopressor administration may increase the stressed volume, eventually increasing cardiac output (CO) and changing RBC velocity [8,9]. Considering that CO is determined by venous return, we hypothesized that the effective circulating blood volume, and thus Pmca, is associated with effective microcirculatory perfusion. This could provide an integrative monitoring tool for assessing hemodynamic coherence and performance, especially when such advanced monitoring methods are not available.

In the present study, we aimed to elucidate our hypothesis in greater depth. To this end, we evaluated the association of Pmca and other determinants of venous return with sublingual microcirculatory variables, RBC velocity, and O_2_ER in anesthetized adults with steady-state physiology. In addition, Computational Fluid Dynamics (CFD) models were developed using clinical data to evaluate the velocity and pressure fields in microvessels.

## 2. MATERIAL AND METHODS

### 2.1 Design

This explorative investigation included individuals who were excluded from a previous prospective observational study due to post-enrollment use of anti-inflammatory medication. The underlying study was conducted in accordance with Good Clinical Practice guidelines, the principles of the Declaration of Helsinki, and relevant regulatory requirements. The original study was registered in ClinicalTrials.gov (NCT03851965, February 22, 2019) [10]. The UHL Institutional Review Board approved the study (IRB no. 60580, 11 Dec 2018), and we obtained written individual informed consent from each participant or next-of-kin. This work is reported according to STROCSS criteria [11].

### 2.2 Study objectives

The goals of the present study were (1) to characterize the relationship between Pmca and sublingual microcirculatory variables and RBC velocity; (2) to characterize the relationship between sublingual RBC velocity and O_2_ER; and (3) to develop CFD models using clinical data to evaluate the velocity and pressure fields in microvessels.

### 2.3 Patient eligibility

We considered adults fulfilling the following criteria: sinus rhythm in electrocardiogram; no evidence of structural heart disease confirmed by preoperative echocardiography; and American Society of Anesthesiologists’ (ASA) physical status I.

### 2.4 Anesthetic management

Before anesthesia induction, all patients received 5 mL kg^-1^ of a balanced crystalloid solution to compensate for preoperative fasting and vasodilation associated with general anesthesia. Anesthesia was induced in the supine position, and included midazolam 0.15-0.35 mg kg^-1^, fentanyl 1 μg kg^-1^, ketamine 0.2 mg kg^-1^, propofol 1.5-2 mg kg^-1^, rocuronium 0.6 mg kg^-1^, and a fraction of inspired oxygen of 0.7. After tracheal intubation, patients were mechanically ventilated using a lung-protective strategy with tidal volume of 7 mL kg^-1^, positive end-expiratory pressure of 6-8 cmH_2_O, and plateau pressure <30 cmH_2_O (Draeger Perseus A500®; Drägerwerk AG & Co., Lübeck, Germany).

General anesthesia was maintained by inhalation of desflurane at an initial 1.0 minimal alveolar concentration. Thereafter, depth of anesthesia was adjusted to maintain Bispectral Index (BIS, Covidien, France) between 40 and 60 [12-14]. Intraoperative fraction of inspired oxygen was then adjusted to maintain an arterial oxygen partial pressure of 80-100 mmHg and normocapnia was maintained by adjusting the respiratory rate as needed [15-17]. Normothermia (37 °C) and normoglycemia were maintained during the perioperative period.

The radial artery was cannulated and connected to a FloTrac/EV1000 clinical platform (Edwards Life Sciences, Irvine, CA, USA) to directly measure mean arterial pressure (MAP), CO and cardiac index (CI), stroke volume (SV), stroke volume variation (SVV), and systemic vascular resistance (SVR). The internal jugular vein was cannulated with a triple-lumen central venous catheter that was connected to a pressure transducer to measure central venous pressure (CVP). Before study measurements, we confirmed that transducers were correctly leveled and zeroed, while the system’s dynamic response was confirmed with fast-flush tests. Artifacts were detected and removed when documented as such and when measurements were out-of-range or systolic and diastolic pressures were similar or abruptly changed (≥40 mmHg decrease or increase within 2 min before and after measurement). Oxygen extraction ratio was calculated as the ratio of oxygen consumption (VO_2_) to oxygen delivery (DO_2_) using the formula O_2_ER = VO_2_ / DO_2_ = (SaO_2_ - ScvO_2_) / SaO_2_.

### 2.5 Calculation of mean circulatory filling pressure analogue and related variables

The methods of the Pmca algorithm have been described in detail before [3,18-20]. Briefly, based on a Guytonian model of the systemic circulation [CO = *V*R = (Pmcf − CVP) / R_VR_], an analogue of Pmcf can be derived using the mathematical model Pmca = (a × CVP) + (b × MAP) + (c × CO) [2,21]. In this formula, *a* and *b* are dimensionless constants (*a* + *b* = 1). Assuming a veno-arterial compliance ratio of 24:1, *a* = 0.96 and *b* = 0.04, reflecting the contribution of venous and arterial compartments, and c resembles arteriovenous resistance and is based on a formula including age, height, and weight [2]:

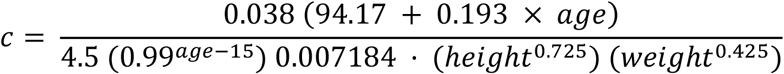

In addition, the following values were determined: (1) Pressure gradient for venous return (PG_VR_) was defined as the pressure difference between Pmca and CVP (PG_VR_ = Pmca – CVP); (2) Resistance to venous return (R_VR_) was defined as the resistance downstream of Pmca to reflect resistance for venous return and was calculated as the ratio of the pressure difference between Pmca and CVP and CO [R_VR_= (Pmca – CVP) / CO]. This formula is used to describe venous return during transient states of imbalances (Pmca is the average pressure in the systemic circulation and R_VR_ is the resistance encountered to the heart) [22,23].

### 2.6 Sublingual microcirculation analysis

Sublingual microcirculation was monitored using SDF+ imaging (Microscan; Microvision Medical BV, Amsterdam, The Netherlands). Microcirculation was assessed 30 minutes after induction of general anesthesia before surgical incision. We recorded sublingual microcirculation videos from at least five sites. All videos were recorded by the same investigator who was blinded to systemic hemodynamic measurements. To optimize video quality, we tried to avoid pressure and movement artefacts, optimized focus and illumination, and cleaned saliva and/or blood from the sublingual mucosa.

### 2.7 Computational Fluid Mechanics

A numerical study was performed utilizing the CFD approach and randomly selected patient data. Randomization was achieved by using random computer-generated numbers. Systemic hemodynamic data were collected by an individual who was blinded to sublingual hemodynamic measurements. The latter were collected by another individual who was blinded to systemic hemodynamic data.

We reconstructed the 2D microvessels and applied CFD to evaluate the correlation of Pmca and RBC velocity with the obtained pressure and velocity fields in microvessels from CFD [i.e., pressure difference (Δp)] under laminar flow assuption. For the blood flow in a blood vessel, Δp is the pressure difference between any two points along its given length, discribing the main driving force of blood motion in the vessel. The discrete equations of fluid flow are the continuity and momentun (Navier-Stokes) equations. These equations form a non-linear system of Partial Differential Equations (PDEs). This system was numerically solved using the Finite Volume (FV) method for the steady state problem. The discretized algebraic system was solved using the Semi-Implicit Method for Pressure Linked Equation algorithm (SIMPLE). The method is described in detail elsewhere [24].

The laminar numerical simulations were performed in the software package Ansys Fluent (Ansys Inc., Canonsburg, PA). Blood was considered as a Newtonian fluid with density ρ = 1050 Kg m^-3^ and kinematic viscosity *v* = 3.2×10-6 m^2^ s^-1^. The numerical scheme was converged when the residuals (errors) of the momentum and continuity equations were less than or equal to the predetermined error, i.e., error =10^−6^, in this study. At the inlet(s), we considered a constant velocity profile obtained from the measurments of this study, specific for each patient. At the outlet(s), a pressure outlet condition was applied, meaning that the pressure at the outlet has a predifined value. Finally, a no slip condition was applied to the microvessel walls, following the rigid wall assumption. More details about the numerical approach can be found elsewhere [25].

### 2.8 Statistical Analysis

Before analysis, all sublingual perfusion videos were evaluated by two experienced raters blinded to all patient data, according to a modified microcirculation image quality score (MIQS) [26]. The best three videos from each recording were analysed offline by a blinded investigator with the AVA4.3C Research Software (Microvision Medical, Amsterdam, the Netherlands) [27,28]. We analysed the De Backer score and De Backer score (small) as density scores, and the Consensus Proportion of Perfused Vessels (Consensus PPV) and Consensus PPV (small) as flow scores. Vessel diameter, vessel length, and RBC velocity were determined with the latest version of AVA Software using a modified optical flow-based algorithm. The method uses per video frame data to measure the overall velocity per vessel segment.

The statistical significance of the hemodynamic variations between the variables analyzed in each microcirculation video was determined by non-parametric ANOVA tests. The studied variables are presented with their mean value (MV) and standard deviation (SD). For correlating the data, the Kendall’s rank correlation between multiple time series was utilized [29]. In this test, we conducted a hypothesis test to determine which correlations are significantly different from zero. The correlation coefficients highlighted in red indicate which pairs of variables have correlations significantly different from zero. Due to the study sample (n=20), post-hoc bootstrapping metrics were used to allow estimation of the sampling distribution using random sampling methods. The analysis, including the post-hoc bootstrapping, was performed in Matlab (MathWorks, Natick, Massachusetts, USA). P values less than 0.05 were deemed significant.

## 3. RESULTS

Twenty patients were included in the study, of whom 12 (60%) were men and 8 (40%) were women, with a median age of 39.5 years (IQR 35.5-44.5). Demographic and clinical characteristics are shown in Supplemental Table 1, while the anesthetic parameters 30 min after induction of anesthesia are depicted in Supplemental Table 2.

### 3.1 Baseline systemic and sublingual microcirculatory variables

Baseline hemodynamic and metabolic parameters were within normal range (Tables 1 and 2). Mean arterial pressure was maintained ≥65 mmHg without vasopressor administration. Sublingual velocity distributions were similar and followed a log-normal distribution, but distinct differences with different mean values were observed from case to case (Figure 1). An additional statistical analysis with a non-parametric ANOVA test showed that the velocity distributions were significantly different among patients (p<0.001). The aforementioned physiological characteristics were translated into a mean DO_2_ and VO_2_ of 973.8±116.2 ml min^-1^ and 247.4±35.6 ml min^-1^, respectively.

**Table 1.** Baseline systemic hemodynamic and metabolic parameters (mean value ± standard deviation)

|  |  |
| --- | --- |
| Heart rate (bmp) | $67.5 \pm 7$ |
| Systolic arterial pressure (mmHg) | $120 \pm 7.4$ |
| Diastolic arterial pressure (mmHg) | $71.3 \pm 7.4$ |
| Mean arterial pressure (mmHg) | $88.1 \pm 7$ |
| Cardiac output (L min<sup>-1</sup>) | $4.8 \pm 1$ |
| Cardiac index (L min<sup>-1</sup> m<sup>-2</sup>) | $2.6 \pm 0.3$ |
| Stroke volume (mL beat<sup>-1</sup>) | $74.7 \pm 9.6$ |
| Stroke volume variation (%) | $5.9 \pm 1.8$ |
| Systemic vascular resistance (dynes sec cm<sup>-5</sup>) | $1306.3 \pm 176.3$ |
| Central venous pressure (mmHg) | $7.1 \pm 0.7$ |
| Analogue of mean circulatory filling pressure (mmHg) | $13.1 \pm 0.9$ |
| Pressure gradient for venous return (mmHg) | $5.9 \pm 0.8$ |
| Resistance to venous return (mmHg min<sup>-1</sup> L<sup>-1</sup>) | $1.2 \pm 0.2$ |
| Oxygen delivery (mL min<sup>-1</sup>) | $973.8 \pm 116.2$ |
| Oxygen consumption (mL min<sup>-1</sup>) | $247.4 \pm 35.6$ |
| Oxygen extraction ratio (%) | $25.8 \pm 2.3$ |
| FiO<sub>2</sub> (%) | $0.3 \pm 0.03$ |
| pH | $7.39 \pm 0.02$ |
| PaO<sub>2</sub> (mmHg) | $92.5 \pm 5.1$ |
| PaCO<sub>2</sub> (mmHg) | $39.2 \pm 1.3$ |
| HCO<sub>3</sub> (mmol L<sup>-1</sup>) | $25.6 \pm 1$ |
| Base deficit (mmol L<sup>-1</sup>) | $2.08 \pm 0.2$ |
| Hemoglobin (g dL<sup>-1</sup>) | $14.1 \pm 0.94$ |
| Glucose (mg dL<sup>-1</sup>) | $113.6 \pm 6.2$ |
| Lactate (mmol L<sup>-1</sup>) | $0.8 \pm 0.2$ |
| SpO<sub>2</sub> (%) | $99.6 \pm 0.5$ |
| SaO<sub>2</sub> (%) | $100 \pm 0.0$ |
| ScvO<sub>2</sub> (%) | $74.2 \pm 2.3$ |
| v-aPCO<sub>2</sub> (mmHg) | $2.8 \pm 0.9$ |

PaO<sub>2</sub>, arterial partial pressure of oxygen; PaCO<sub>2</sub>, arterial partial pressure of carbon dioxide; SpO<sub>2</sub> = oxygen saturation of hemoglobin; ScvO<sub>2</sub>, Central venous oxygen saturation; v-aPCO<sub>2</sub>, venous-to-arterial carbon dioxide difference.

**Table 2.** Baseline values of microcirculatory variables (mean value ± standard deviation)

|  |  |
| --- | --- |
| De Backer score ($\text{mm}^{-1}$) | $3.7 \pm 1.2$ |
| De Backer score (small) ($\text{mm}^{-1}$) | $2 \pm 1.1$ |
| Consensus proportion of perfused vessels (%) | $94.2 \pm 5.7$ |
| Consensus proportion of perfused vessels (small) (%) | $88.2 \pm 10$ |
| Vessel length ($\mu\text{m}$) | $137.3 \pm 96.8$ |
| Vessel diameter ($\mu\text{m}$) | $17.2 \pm 4$ |
| Velocity of red blood cells ($\mu\text{m s}^{-1}$) | $14.7 \pm 9$ |

**Figure 1.**
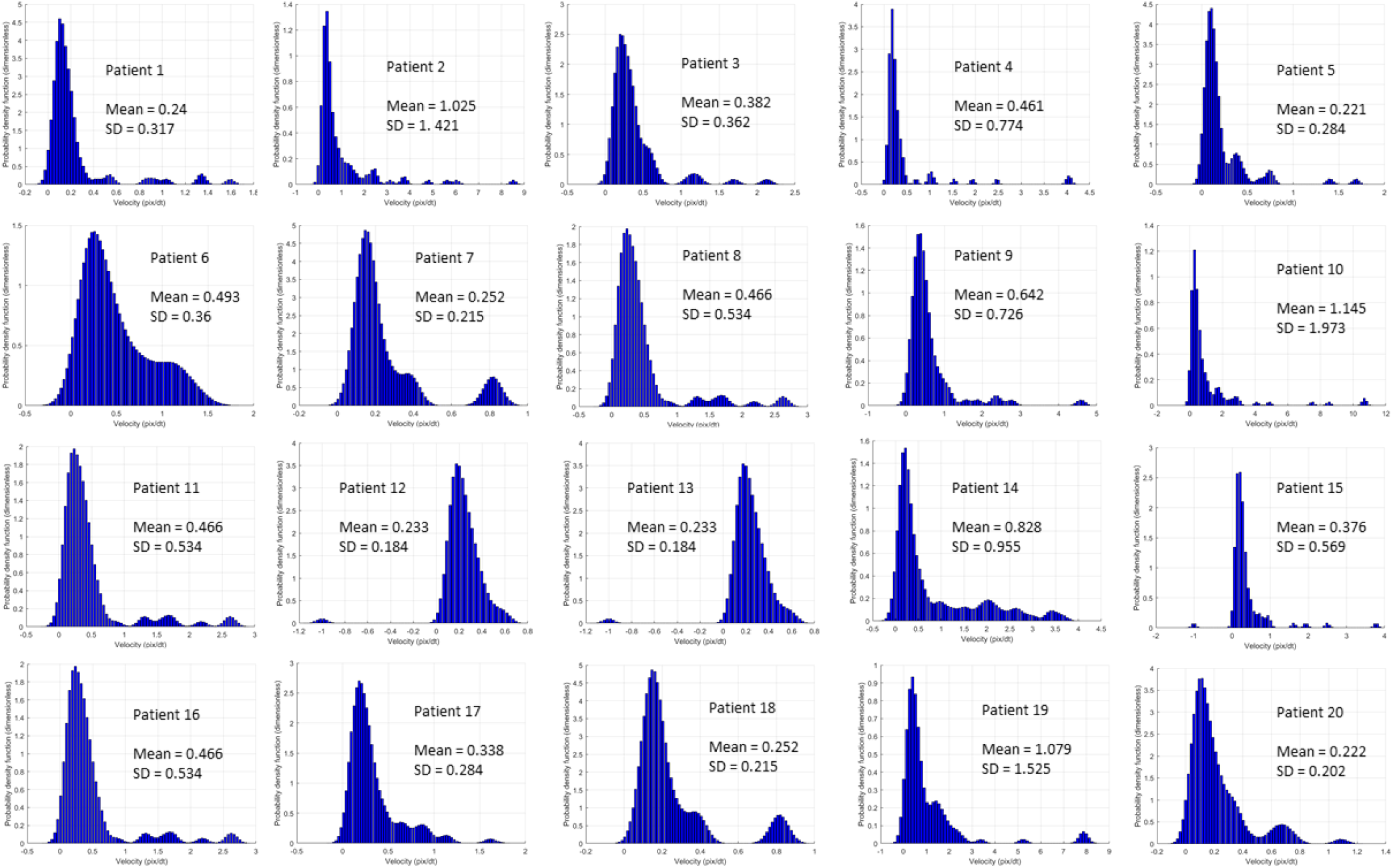
Probability density functions of the velocity distributions. Note the velocity values (mean) and the standard deviation (SD).

### 3.2 Correlation of Pmca with systemic and microcirculatory variables

The correlation of Pmca with systemic hemodynamic variables and sublingual microcirculatory flow and density variables is depicted in Table 3.

**Table 3.** Correlation of Pmca with systemic hemodynamic variables and sublingual flow and density variables.

| **Systemic hemodynamic variables** | **Spearman's rho** | **p-value** |
| --- | --- | --- |
| Cardiac output ($\text{L min}^{-1}$) | 0.173 | 0.31 |
| Cardiac index ($\text{L min}^{-1} \text{m}^{-2}$) | 0.145 | 0.41 |
| Mean arterial pressure (mmHg) | 0.398 | 0.012 |
| Systemic vascular resistance ($\text{dynes sec cm}^{-5}$) | 0.058 | 0.75 |
| Pressure gradient for venous return (mmHg) | 0.438 | 0.008 |
| Resistance of venous return ($\text{mmHg min}^{-1} \text{L}^{-1}$) | 0.203 | 0.23 |
| **Sublingual microcirculatory flow and density variables** | **Spearman's rho** | **p-value** |
| De Backer score ($\text{mm}^{-1}$) | -0.189 | 0.27 |
| De Backer score (small) ($\text{mm}^{-1}$) | 0.011 | 0.97 |
| Consensus PPV (%) | 0.017 | 0.95 |
| Consensus PPV (small) (%) | 0.06 | 0.74 |

### 3.3 Correlation of Pmca with sublingual RBC velocity and microvessel length

A negative correlation was observed between Pmca and RBC velocity (r=-0.03, p=0.87). Interestingly, a constant Pmca value of 14 mmHg was observed in all individuals with sublingual RBC velocity of 6-24 μm s^-1^. On the contrary, a Pmca <14 mmHg was observed in those with RBC velocity >24 μm s^-1^ (Figure 2). In addition, a positive correlation between Pmca and sublingual microvessel length was also observed (r=0.04, p=0.82; Supplementary Figure 1).

**Figure 2.**
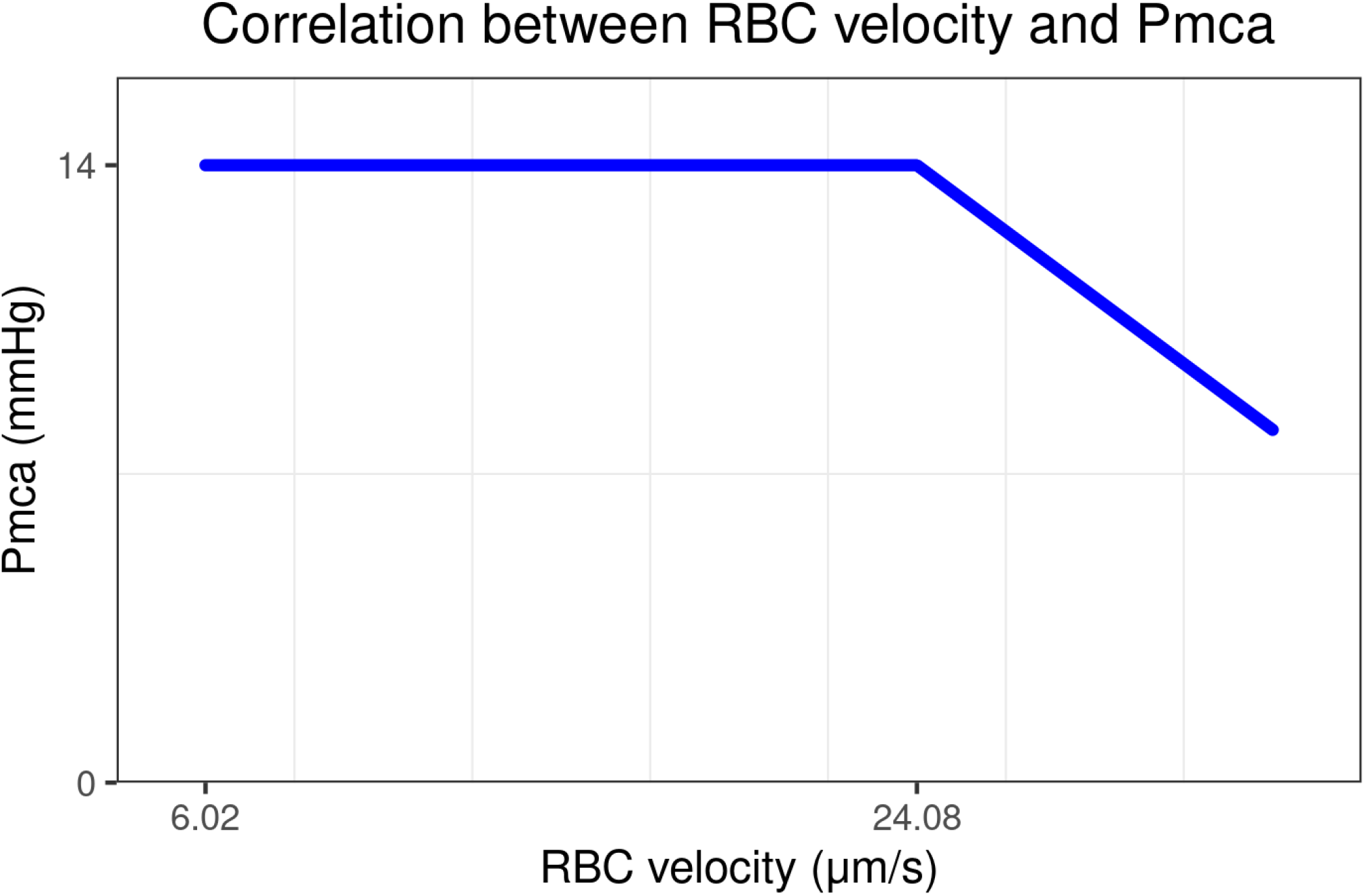
Correlation of Pmca with sublingual RBC velocity in individuals with steady-state physiology, effective coupling between the macro- and microcirculation, and normal tissue oxygen extraction ratio. A constant Pmca of 14 mmHg was observed in individuals with RBC velocity of 6-24 μm/s, while a Pmca of <14 mmHg was observed in those with RBC velocity >24 μm/s. Pmca, mean circulatory filling pressure analogue; RBC, red blood cell.

### 3.4 Correlation of sublingual RBC velocity with microvessel diameter and length

A negative correlation between RBC velocity and microvessel length (r=-0.19, p=0.27) was observed in all patients. Additionally, a positive correlation was observed between RBC velocity and microvessel diameter (r=0.68, p<0.001; Supplementary Figure 2).

### 3.5 Correlation of sublingual RBC velocity with density and flow variables and O_2_ER

A positive correlation was observed between RBC velocity and De Backer score (r=0.2, p=0.24), De Backer score (small) (r=0.26, p=0.125), and Consensus PPV (small) (r=0.07, p=0.73) (Supplementary Table 3, Supplementary Figure 3). Also, a positive correlation was observed between mean RBC velocity and O_2_ER (r=0.034, p=0.87).

### 3.6 Computational Fluid Mechanics

Reconstruction of the 2D microvessel and application of CFD to evaluate the velocity and pressure fields in microvessels are depicted in Supplementary Figures 4-12. Correlation of Pmca with RBC velocity and pressure drop (Δp) is depicted in Figure 3. Interestingly, when Pmca ranged between 11 mmHg and 15 mmHg, Δp fluctuated between 0.02 Pa and 0.1 Pa. Bootstrapping metrics (n=30) revealed a statistically significant negative correlation between Pmca and Δp (r=-0.30, p=0.02) (Supplementary Figure 13).

**Figure 3.**
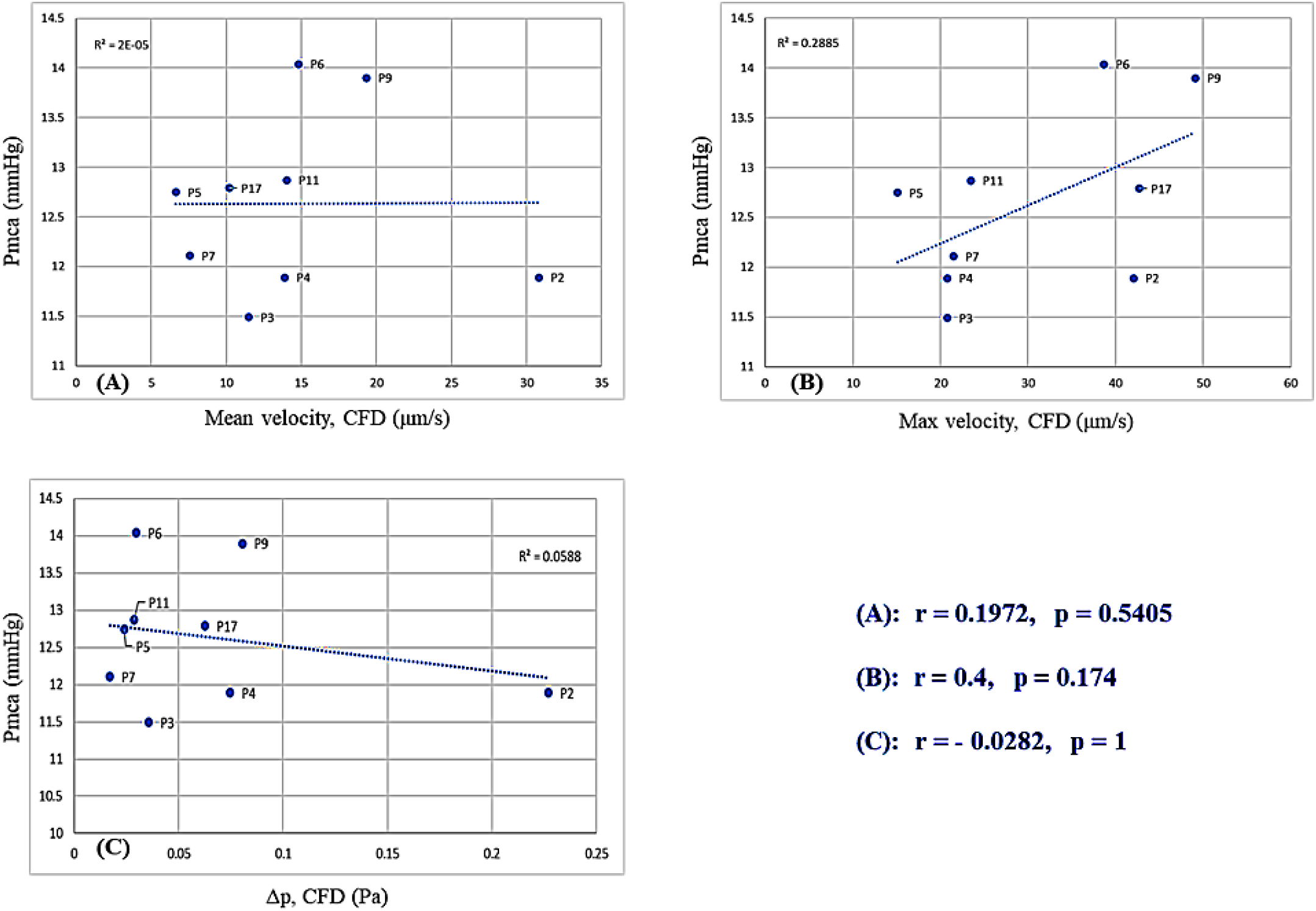
Computational fluid dynamics simulation: (A) Correlation between Pmca and mean RBC velocity; (B) Correlation between Pmca and maximum velocity in the domain; (C) Correlation between Pmca and pressure difference (Δp).

## 4. DISCUSSION

In this explorative study with anesthetized individuals in steady-state physiology, a constant Pmca value of 14 mmHg was observed in those with sublingual RBC velocity of 6-24 μm s^-1^, while a Pmca <14 mmHg was observed in those with RBC velocity >24 μm s^-1^. A positive correlation was observed between RBC velocity and O_2_ER. In addition, CFD modeling simulation revealed a negative correlation between Pmca και Δp. When Pmca ranged between 11 mmHg and 15 mmHg, Δp constantly fluctuated between 0.02 Pa and 0.1 Pa. The present study provides a novel method to monitor hemodynamic coherence and tissue perfusion, which can aid in the identification of novel hemodynamic phenotypes and enhance microcirculation-guided therapeutic strategies, optimizing local delivery of oxygen.

An intact coherence between the macro- and microcirculation facilitates DO_2_ to the parenchymal cells. Patients undergoing general anesthesia and critically ill patients may experience a transient or prolonged loss of this coherence, which may lead to tissue hypoperfusion and organ injury. Although the expansion of hemodynamic monitoring to include monitoring of the microcirculation may be helpful in guiding management [30], its visualization and assessment remains technically challenging. Therefore, identifying potential systemic hemodynamic variables that would enable prediction of microcirculatory behavior is of highest interest [31]. In this clinical study with CFD analysis, we report that a Pmca of 14 mmHg is associated with physiological hemodynamic coherence, effective microcirculatory perfusion, and oxygen transport to tissue. As hemodynamic reference values or thresholds are needed to define microcirculatory alterations as persisting, Pmca may prove an indirect assessment method of tissue perfusion, allowing microcirculation-guided resuscitation and aiding in the identification of novel hemodynamic phenotypes [5,28].

In normal conditions, organ perfusion is dependent upon CO and the vascular resistance across an organ, while in circulatory shock, fluid resuscitation is often necessary to achieve maximal tissue RBC perfusion [32]. The present study included individuals with steady-state physiology, intact vascular regulation, and effective coupling between the macro- and microcirculation, which allowed the description of their functional state. Of note, the association between systemic hemodynamic and microcirculatory variables may still exist when hemodynamic coherence is lost [31]. This is important for the monitoring and treatment of hemodynamic abnormalities, especially in the early phase of diseases during which hemodynamic coherence is usually maintained [31]. In our patients with dynamic hemodynamic equilibrium, a lower Pmca value (<14 mmHg) was correlated with higher RBC velocity, probably due to an increase in vascular capacitance and sublingual vessel diameter and/or CO. However, there must be always a limit under which decreases in Pmca and venous return impair microcirculatory blood flow [5,33]. On the contrary, a higher Pmca may be the result of excessive vasoconstriction, eventually resulting in hemodynamic incoherence and tissue hypoperfusion. These may explain the detrimental effects of higher doses of adrenergic vasoconstrictive agents and support the recent trend towards a perfusion-centered resuscitation strategy instead of standard pressure-guided treatment [34,35].

In the present study, mean RBC velocity was 15±9 μm s^-1^, which is significantly lower compared to this reported in other studies including healthy individuals. Edul et al. reported a normal RBC velocity of 1331±90 μm s^-1^ [8,9], while Rovas et al. recently reported a normal RBC velocity of approximately 102 μm s^-1^ [36]. Nevertheless, the available evidence is not sufficient to rule out different RBC velocities in healthy people [37]. The lower RBC velocity in our patients is suggestive of a hypodynamic microcirculatory state, with the normal O_2_ER indicating a physiological balance between DO_2_ and VO_2_. This, together with the microcirculatory density and flow scores in the present study, strengthen the evidence revealing that tissue oxygenation can be maintained in very low RBC velocities. Theoretically, oxygen transport to tissue is facilitated when RBC velocity is low, while tissue hypoxia may develop when RBC velocity increases because the capillary transit time of the RBCs may not be sufficient to unload oxygen completely [6,8]. However, whether a hyperdynamic microcirculatory flow is always associated with tissue hypoxia remains controversial. In septic patients, individual changes in sublingual RBC velocity have been correlated with those in cardiac index after a fluid bolus, but in the face of an unchanged perfused vascular density [9]. In others, a fluid challenge may improve O_2_ER by increasing Pmcf and venous return [38]. The aforementioned data and the findings of the present study encourage further translational research aimed at the elucidation of their broader implications.

The application of CFD simulations revealed a negative correlation between Pmca και Δp. For the blood flow in a vessel or organ, Δp is the pressure difference between any two points along a given length of the vessel or the difference between the arterial and venous pressures, respectively. In our CFD analysis with laminar flow conditions, with the vascular resistance being independent of flow and pressure, an increase in resistance would decrease flow at any given Δp. In clinical practice, a fluid challenge may increase the stressed volume (and thus Pmca and CO) until a certain point, but may not always improve microcirculatory perfusion. On the other hand, fluid overload increases CVP, which decreases venous return and retrogradely increases post-capillary venular pressure, impairing microcirculatory perfusion [39-41]. The association between the Pmca and Δp ranges in our study (Figure 3) further enhances the potential of Pmca to serve as a hemodynamic coherence monitoring tool. Bedside estimation of Pmca can track the effective circulatory blood volume and a constant value of 14 mmHg may ensure an adequate balance between macrocirculatory pressure and microcirculatory perfusion [42,43]. The post-hoc bootstrapping metrics in the present study strongly encourage the evaluation of our findings in larger studies.

To the best of our knowledge, this is the first report of the association between Pmcf/Pmca and sublingual RBC velocity and Δp. Another strength is that data analyses were performed by blinded investigators, thus preventing inter-observer bias and increasing the credibility of study conclusions. Although the present study includes a small patient sample, bootstrapping metrics revealed a statistically significant negative correlation between Pmca and Δp. Mean age in our patients was 39.5 years and the results of the present analysis may be different in older individuals. In addition, anesthesia can lower resting metabolic rate and reduce global VO_2_, and has been associated with a reduction in tissues ability to extract oxygen. In the present study, however, we used desflurane for maintenance because it produces mild and stable effects on the microcirculation compared to other agents [12].

## 5. CONCLUSIONS

The intact regulatory mechanisms may maintain a physiological coupling between systemic hemodynamics and tissue perfusion and oxygenation when Pmca is 14 mmHg, indicating its potential to serve as a marker of hemodynamic coherence.

## Data Availability

Data can be made available upon request after publication through a collaborative process. Researchers should provide a methodically sound proposal with specific objectives in an approval proposal. Please contact the corresponding author for additional information.

## Acknowledgements

This research was partially supported by the Hellenic Society of Cardiopulmonary Resuscitation, Athens, Greece. The authors would like to thank the medical and nursing stuff of the Department of Anesthesiology of the University Hospital of Larisa, Greece, for their assistance during the study period. We are also thankful to Z. Hossain, medical software engineer at Microvision Medical (Amsterdam, The Netherlands), who provided expertise that greatly assisted the research.

## Authors’ contributions

Conceptualization: AC. Data curation: AC, MX. Formal analysis: AC, MX. Methodology: AC, MX. Project administration: AC. Visualization: AC, MX. Writing - original draft: AC. Writing - review & editing: AC, MX.

