## Supplemental Material for "Association of effective circulating blood volume with sublingual RBC velocity and microvessel pressure difference in anesthetized individuals: A clinical investigation and computational fluid dynamics modeling"

**Supplemental Figure 1**

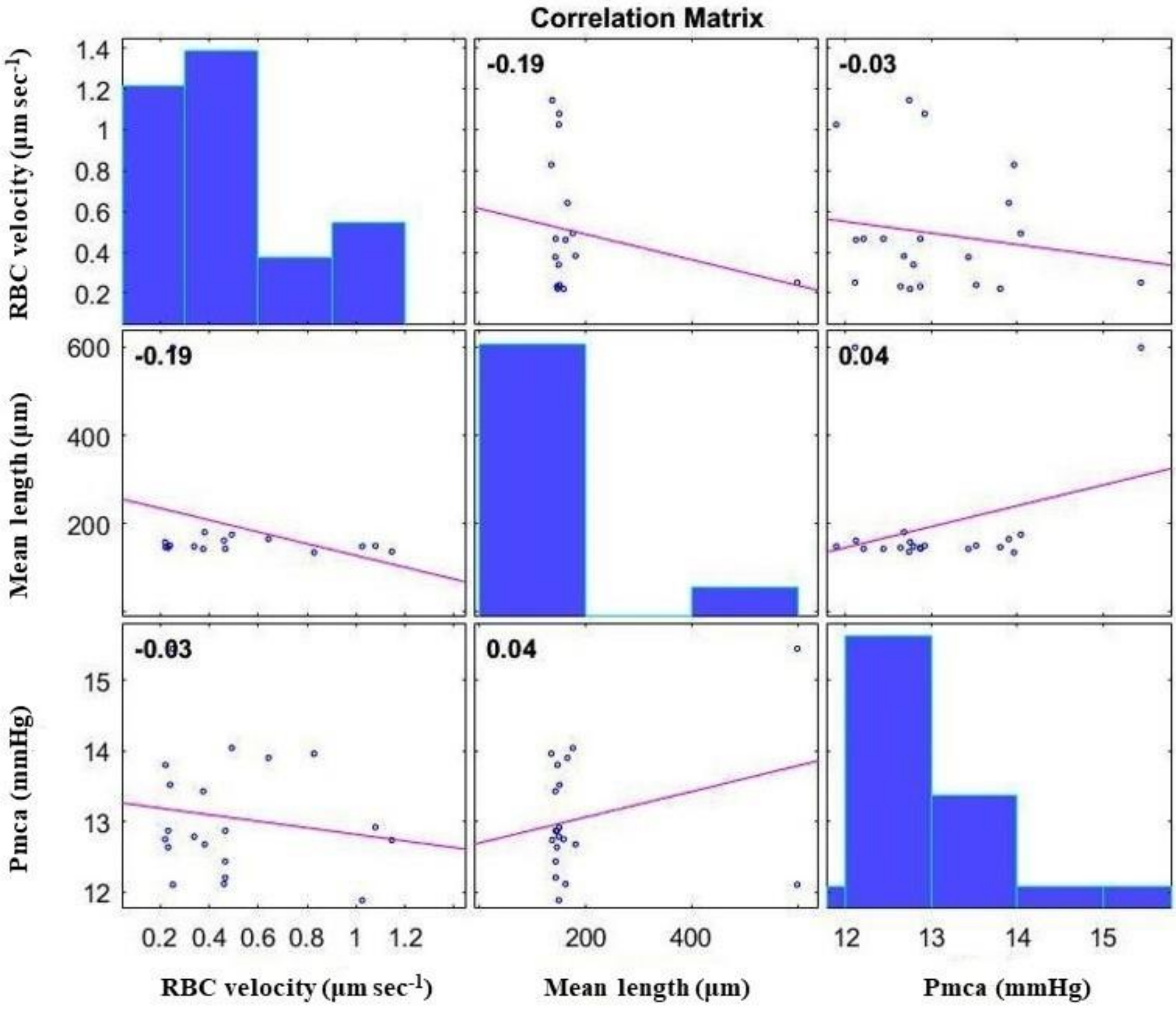


**Supplemental Figure 1.** Correlations between Pmca, microvessel length, and RBC velocity.

**Supplemental Figure 2**

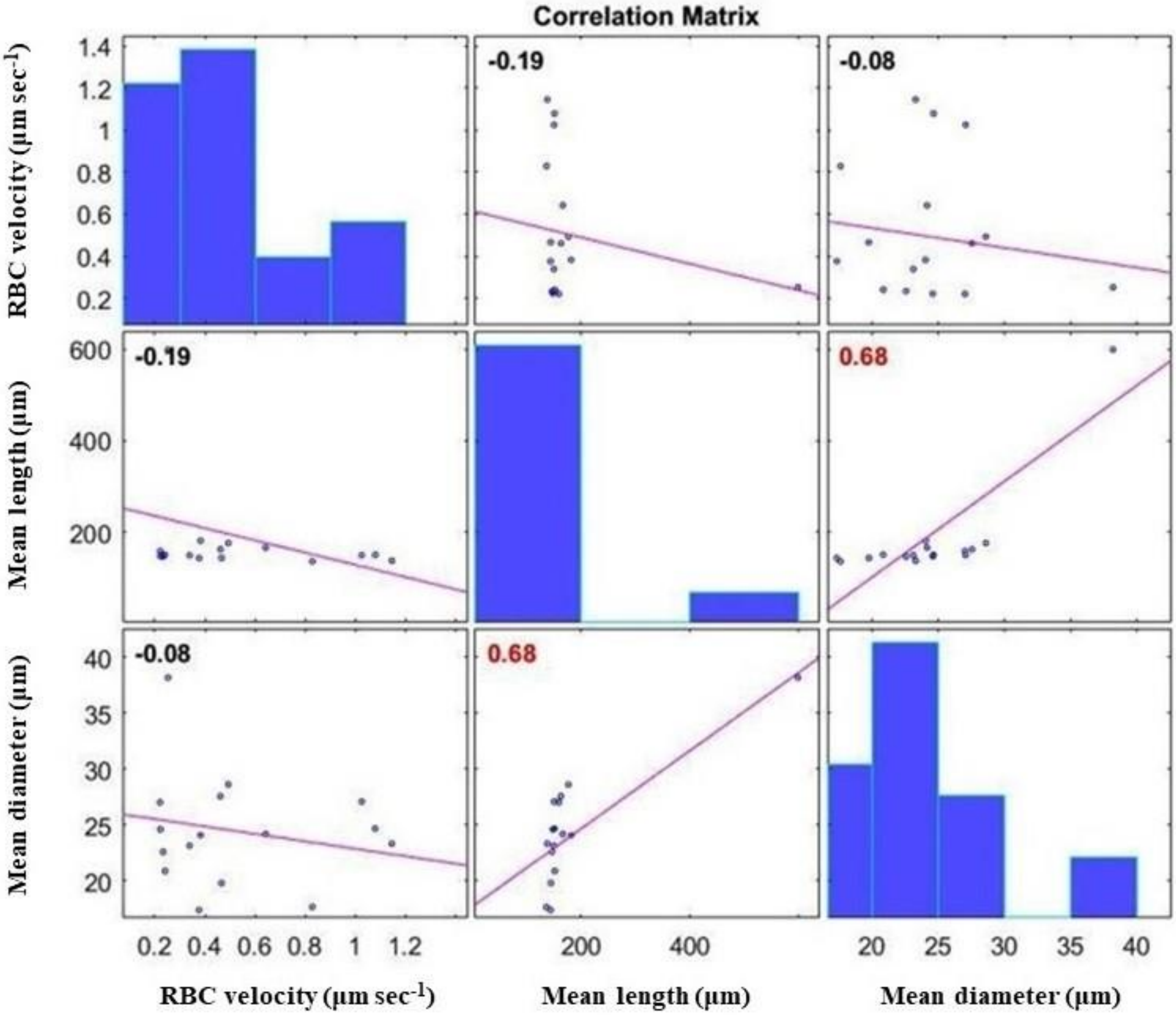


**Supplemental Figure 2.** Correlations of RBC velocity with microvessel length and diameter.

**Supplemental Figure 3**

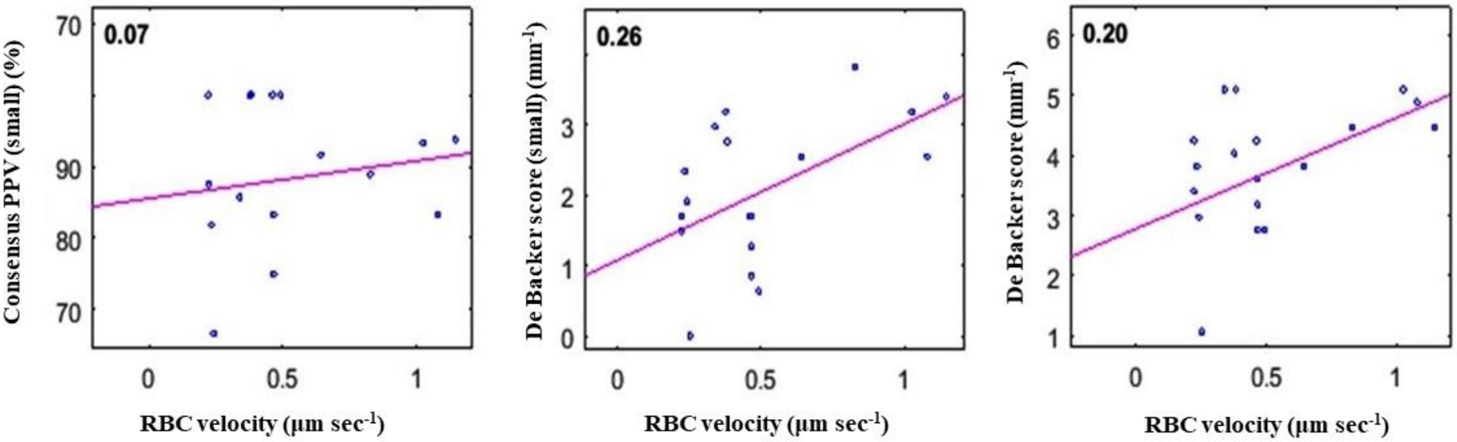


**Supplemental Figure 3.** Correlation of sublingual RBC velocity with Consensus PPV (small), De Backer score (small), and De Backer score.

**Supplemental Figure 4**

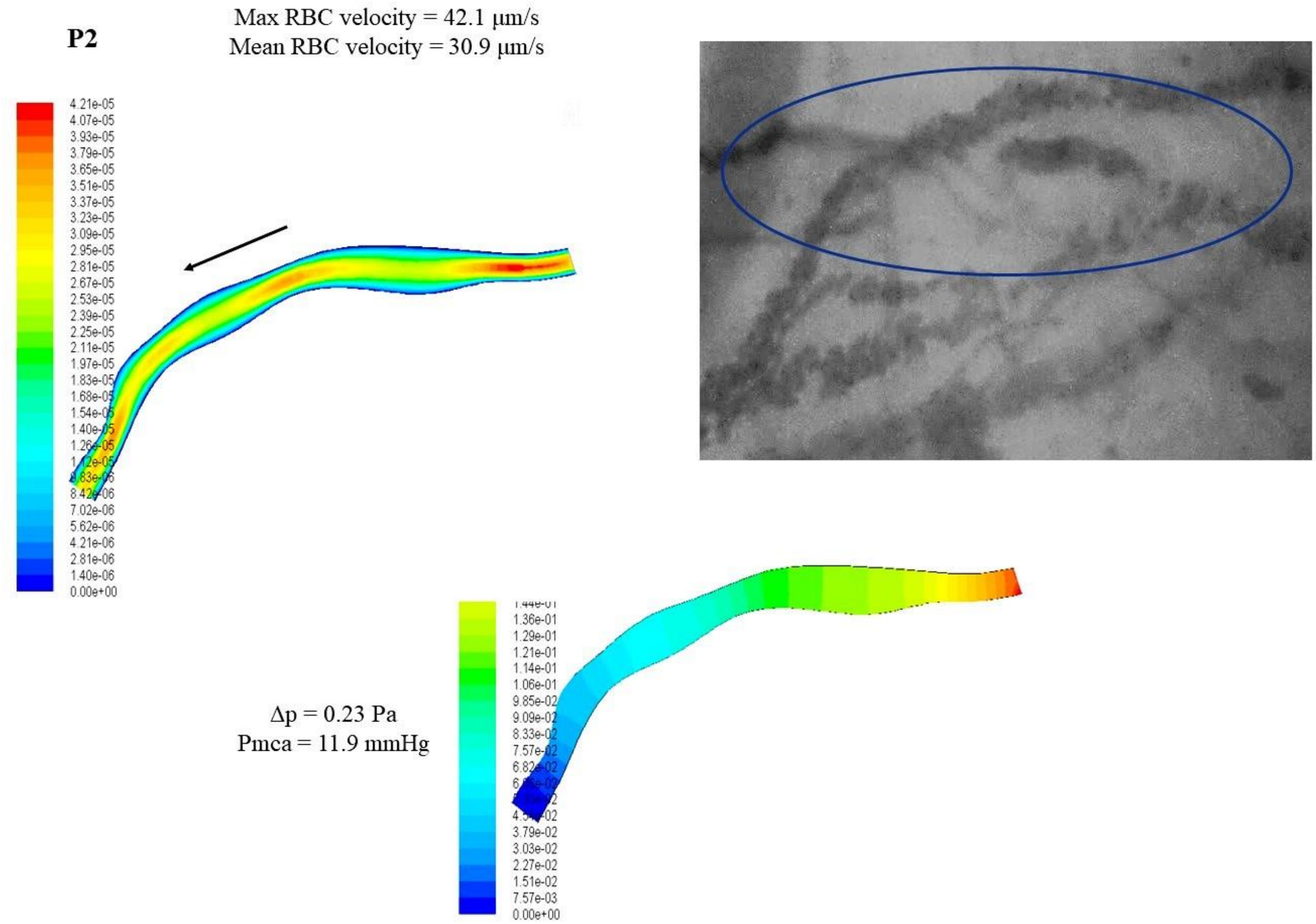


**Supplemental Figure 4.** Reconstruction of the 2D microvessel and application of CFD to evaluate the velocity and pressure fields in microvessels.

**Supplemental Figure 5**

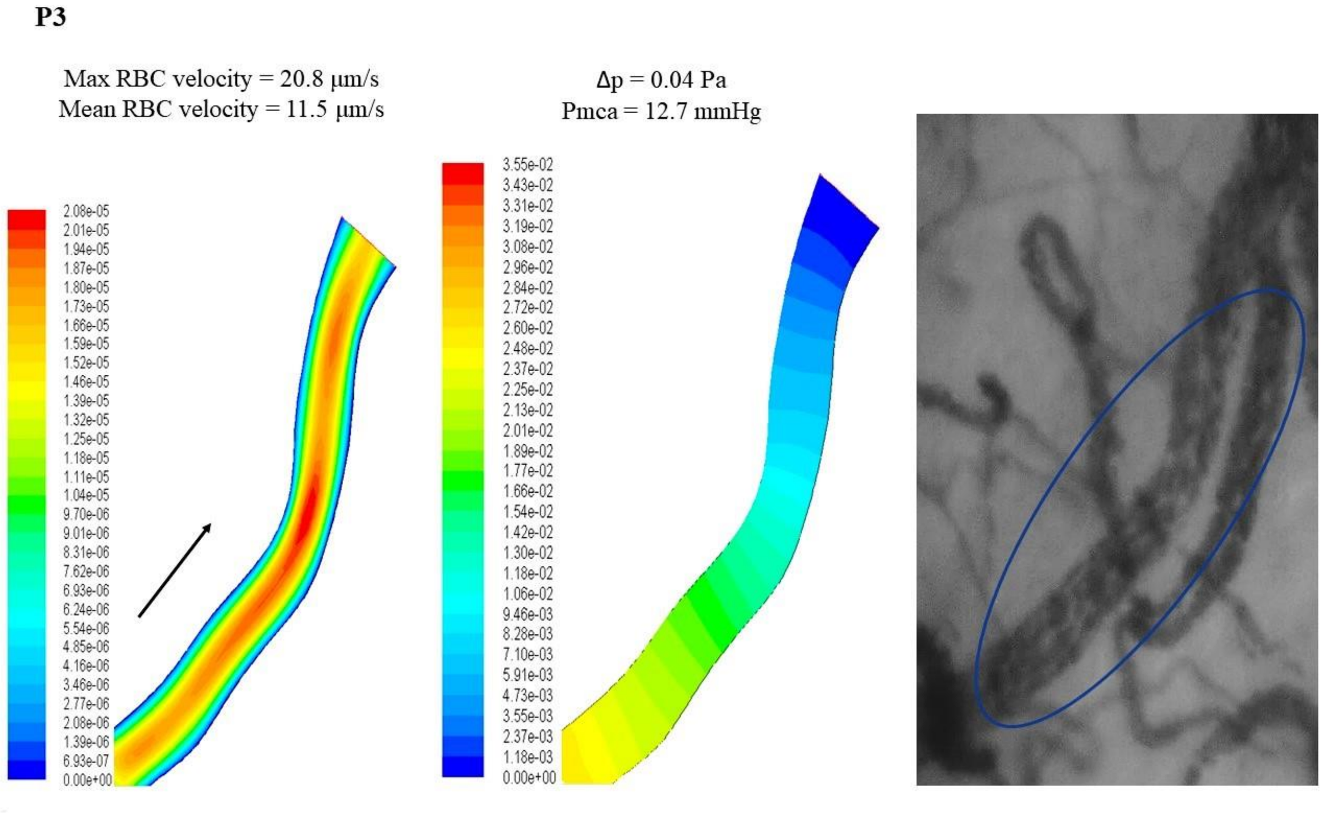


**Supplemental Figure 5.** Reconstruction of the 2D microvessel and application of CFD to evaluate the velocity and pressure fields in microvessels.

**Supplemental Figure 6**

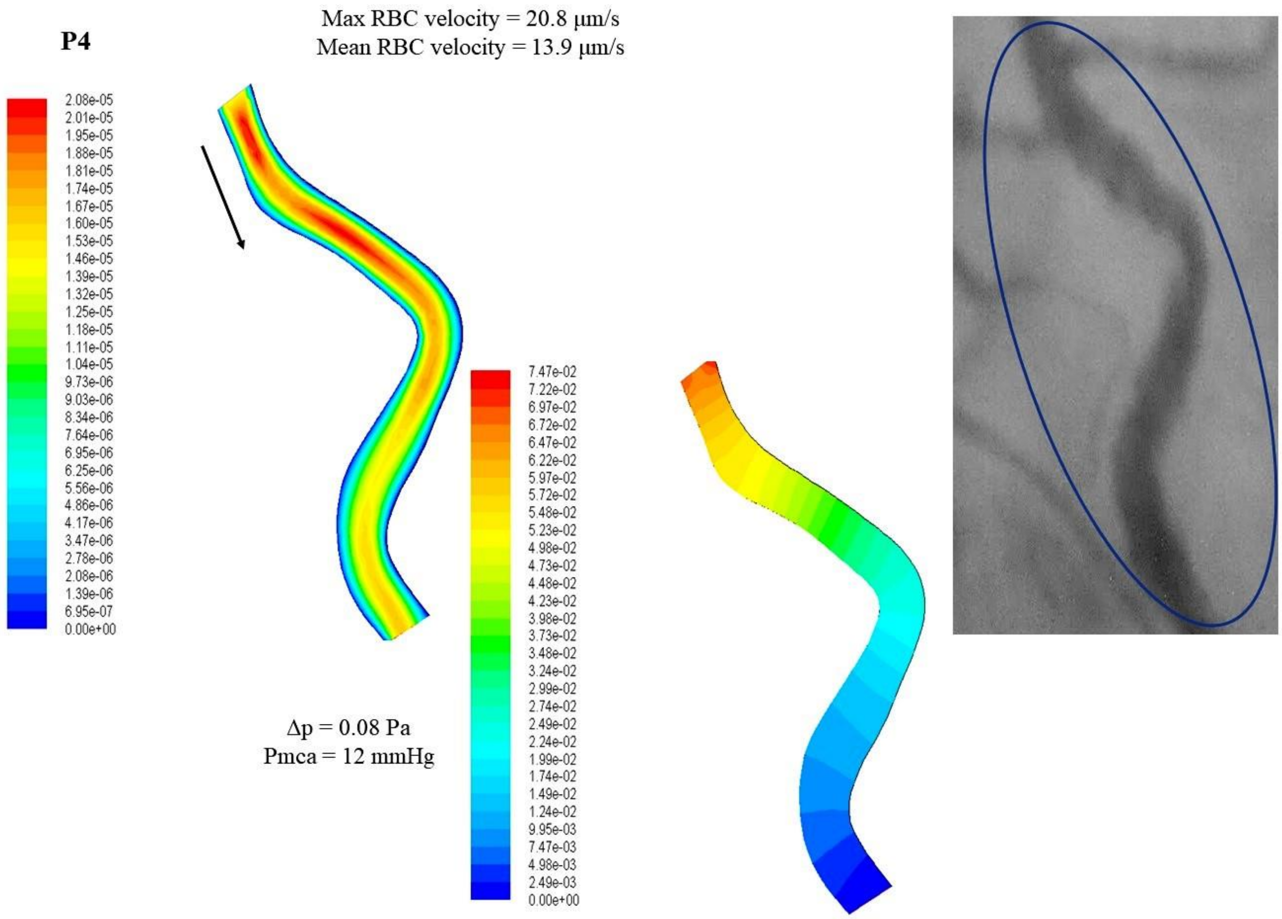


**Supplemental Figure 6.** Reconstruction of the 2D microvessel and application of CFD to evaluate the velocity and pressure fields in microvessels.

**Supplemental Figure 7**

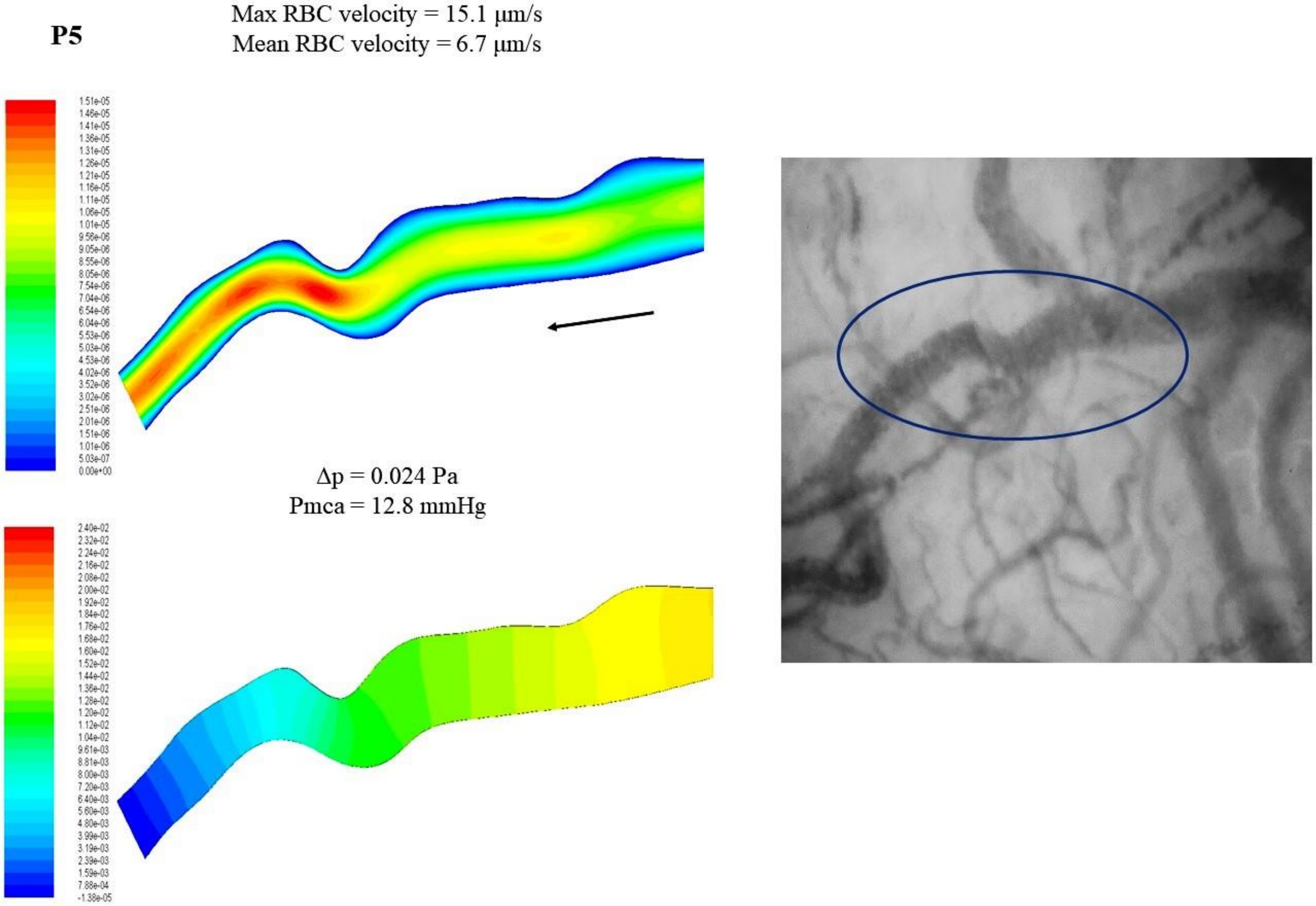


**Supplemental Figure 7.** Reconstruction of the 2D microvessel and application of CFD to evaluate the velocity and pressure fields in microvessels.

**Supplemental Figure 8**

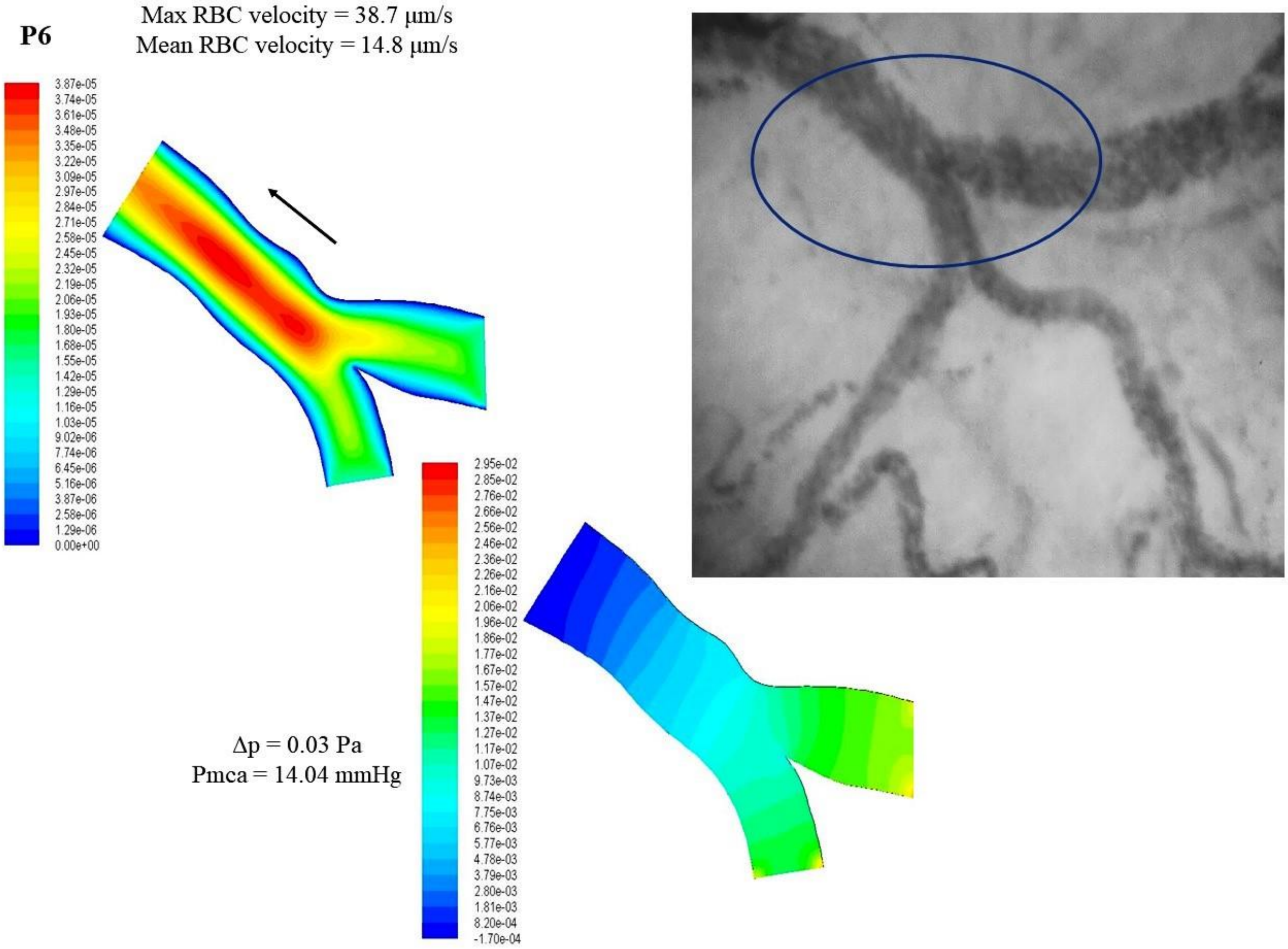


**Supplemental Figure 8.** Reconstruction of the 2D microvessel and application of CFD to evaluate the velocity and pressure fields in microvessels.

**Supplemental Figure 9**

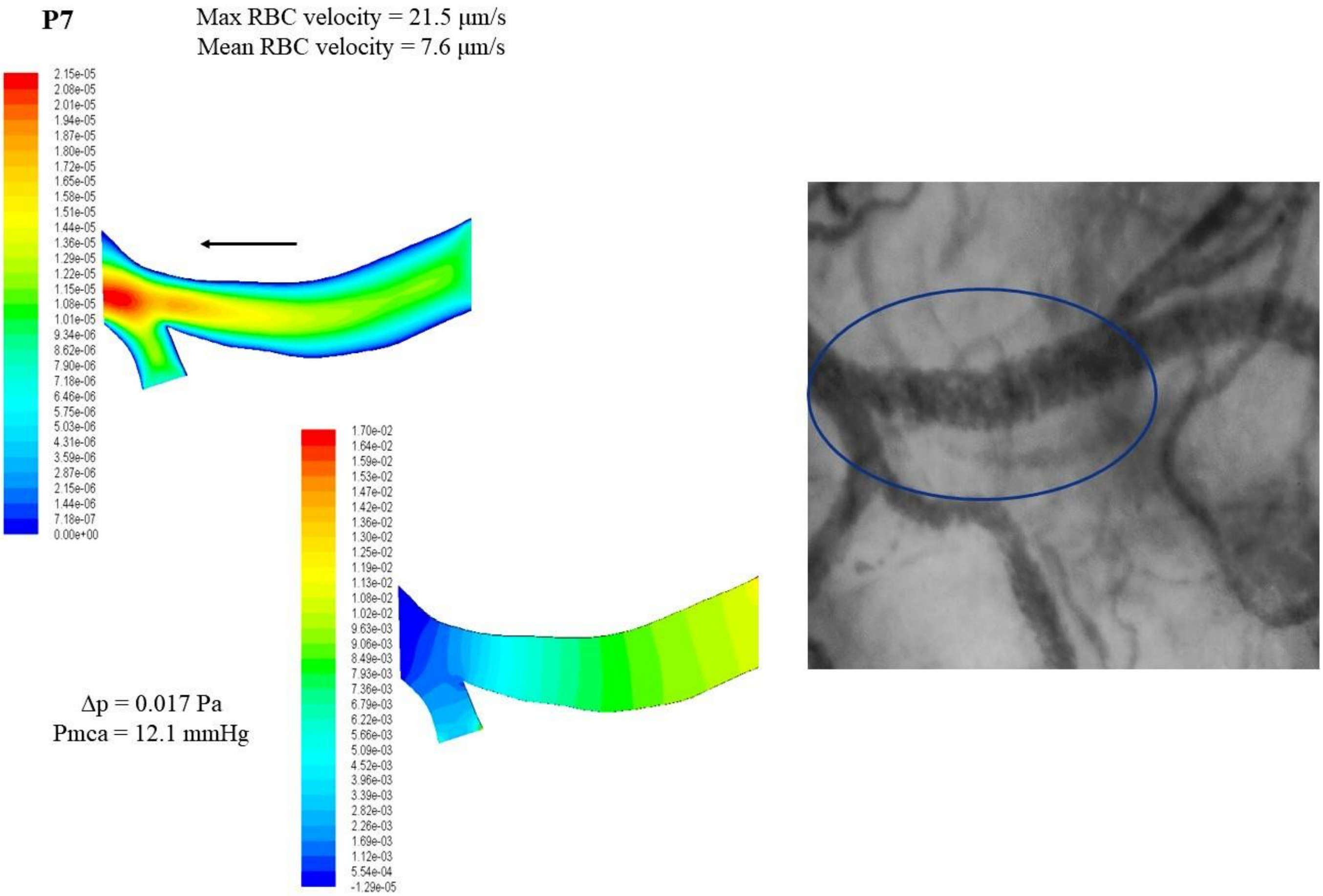


**Supplemental Figure 9.** Reconstruction of the 2D microvessel and application of CFD to evaluate the velocity and pressure fields in microvessels.

**Supplemental Figure 10**

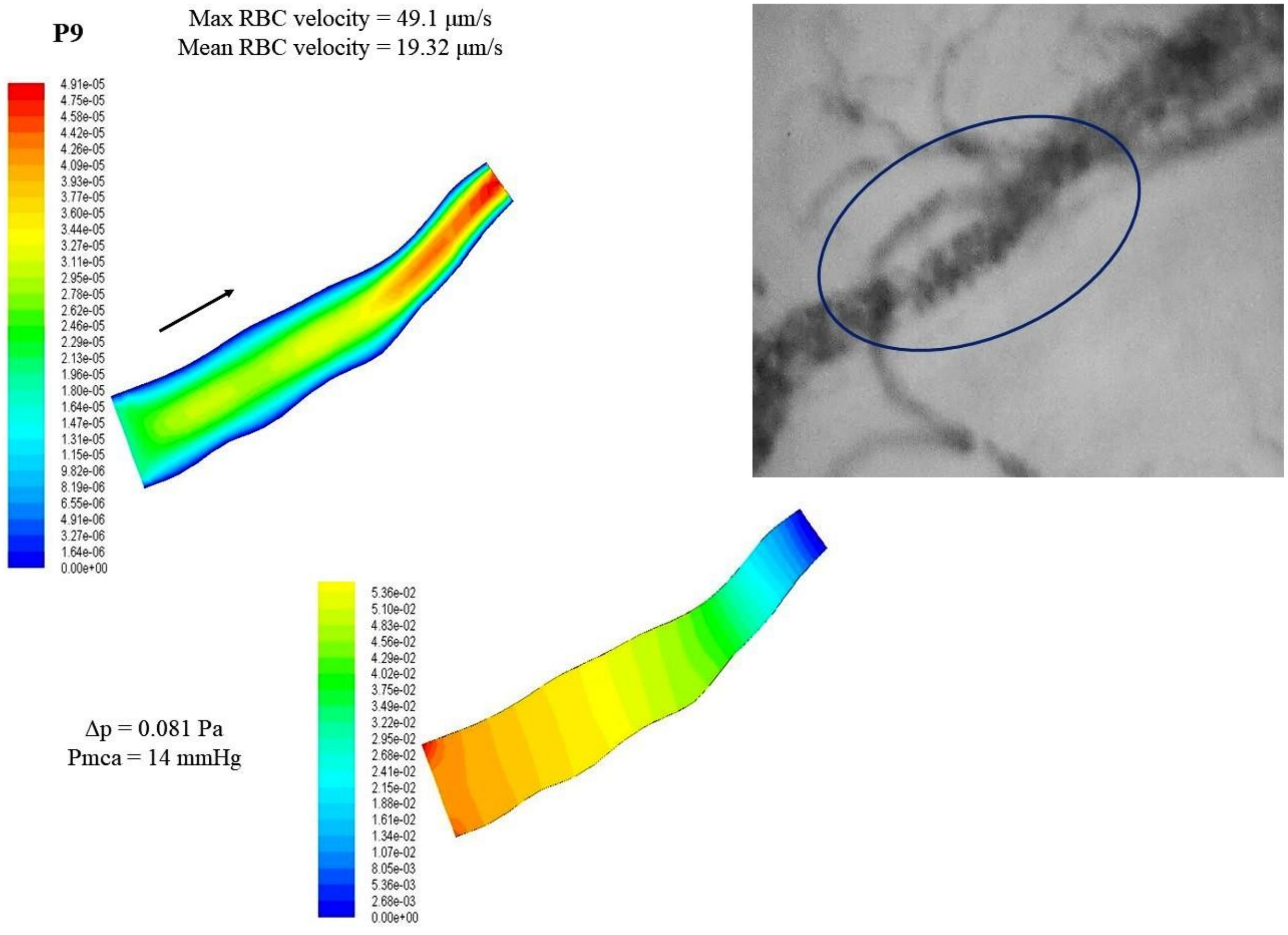


**Supplemental Figure 10.** Reconstruction of the 2D microvessel and application of CFD to evaluate the velocity and pressure fields in microvessels.

**Supplemental Figure 11**

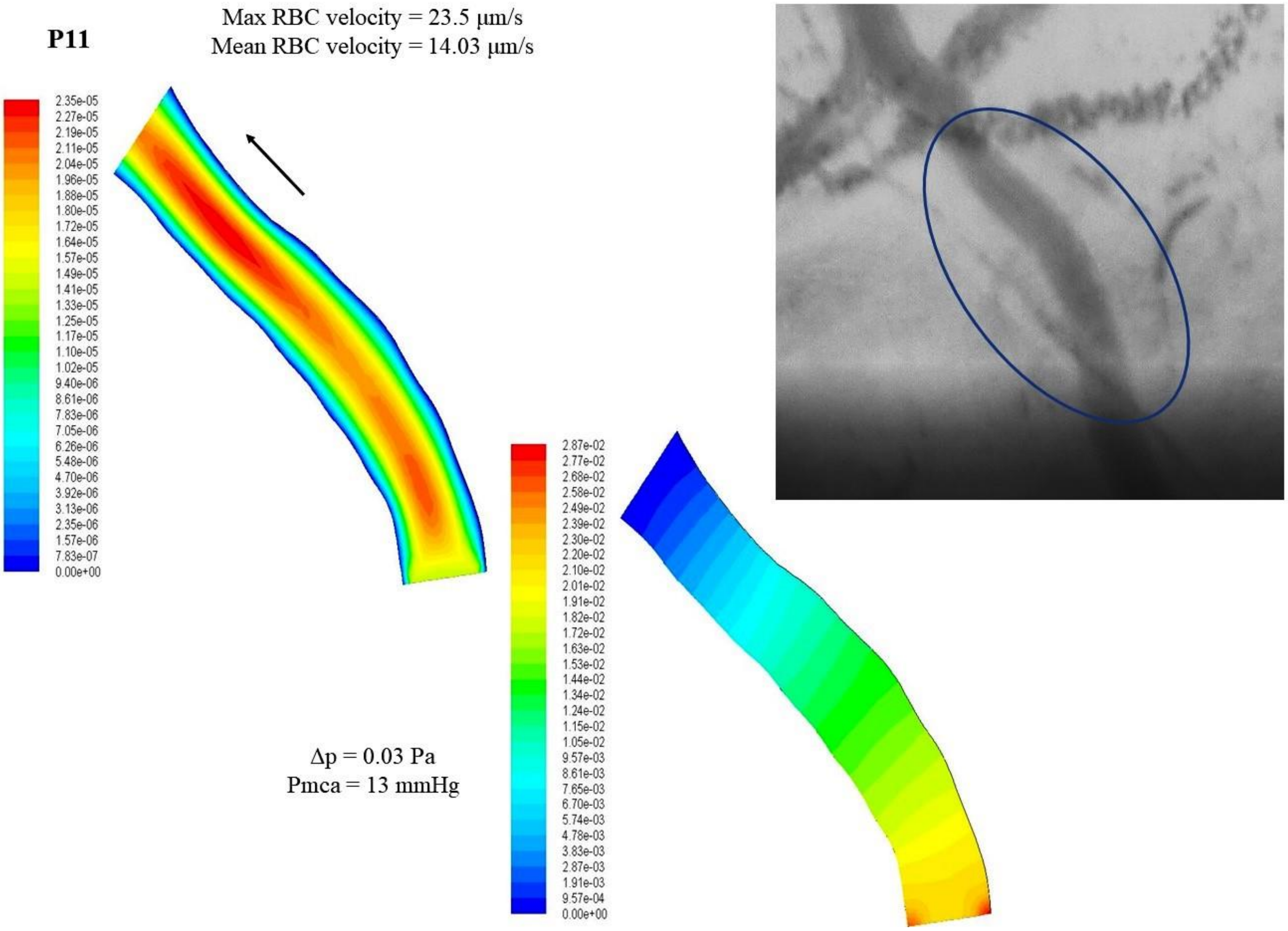


**Supplemental Figure 11.** Reconstruction of the 2D microvessel and application of CFD to evaluate the velocity and pressure fields in microvessels.

**Supplemental Figure 12**

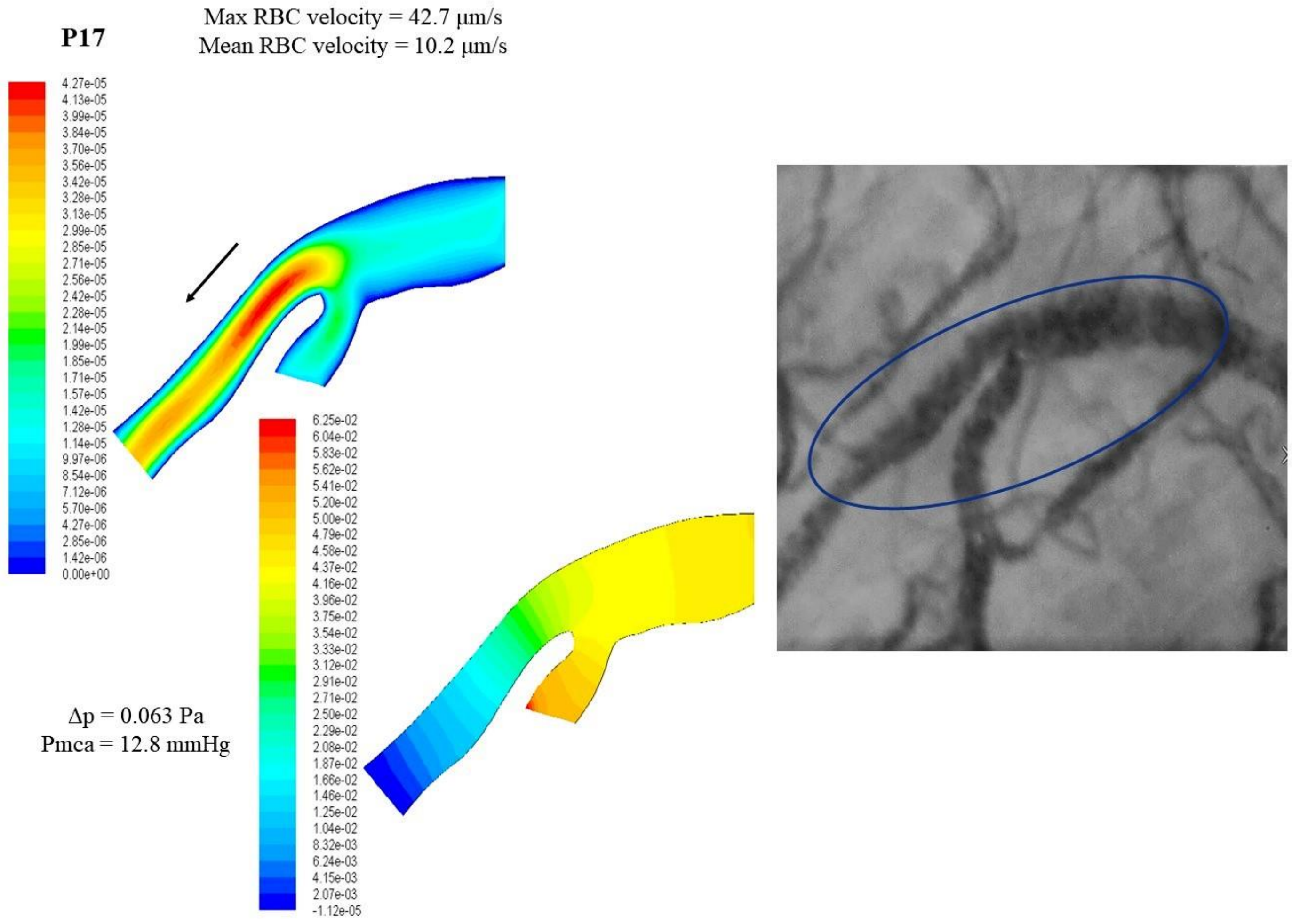


**Supplemental Figure 12.** Reconstruction of the 2D microvessel and application of CFD to evaluate the velocity and pressure fields in microvessels.

**Supplemental Figure 13**

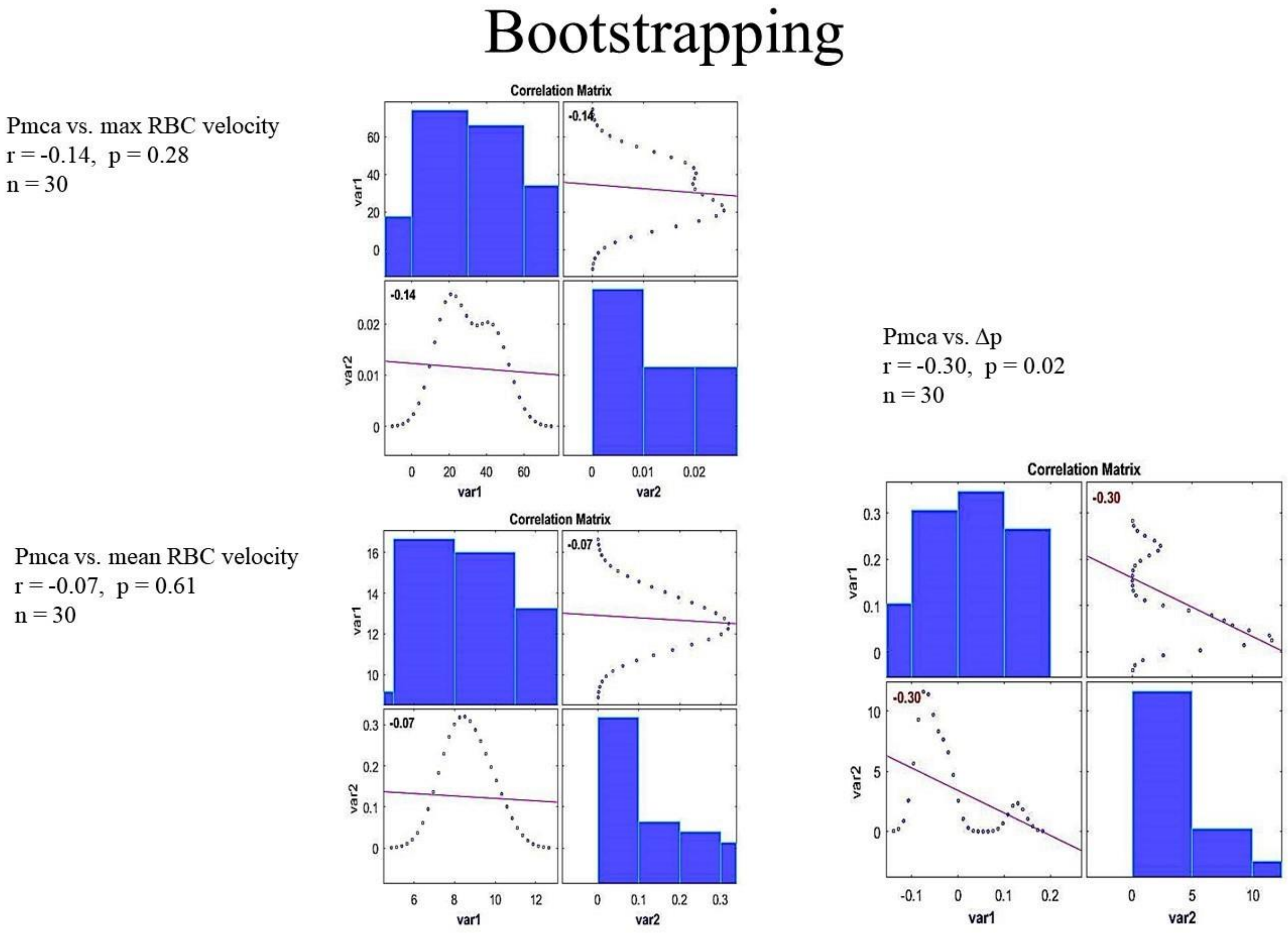


**Supplemental Figure 13.** Bootstrapping metrics analysis (n=30) investigating the correlation between Pmca and Δp.

**Supplemental Table 1. Baseline characteristics of the patients**

| | |
|---|---|
| Age (years), median (IQR) | 39.5 (35.5-44.5) |
| Sex - male, n (%) | 12 (60) |
| Sex - female, n (%) | 8 (40) |
| BMI (kg/m$^2$), median (IQR) | 25.1 (23.15-25.8) |
| Body Surface Area (m$^2$), median (IQR) | 1.94 (1.83-2.02) |
| Type of planned surgery | |
| Gastrointestinal, n (%) | 7 (35) |
| Gynecological, n (%) | 3 (15) |
| Thyroidectomy, n (%) | 2 (10) |
| Urological, n (%) | 8 (40) |

**Supplemental Table 2. Baseline anesthetic parameters 30 min after induction of anesthesia (mean value ± standard deviation)**

| | |
|---|---|
| Bispectral index | 42.5 ± 1.5 |
| Temperature (°C) | 36.7 ± 0.1 |
| Tidal volume (ml $kg^{-1}$ IBW) | 560 ± 52.2 |
| Respiratory rate ($min^{-1}$) | 13.7 ± 1.1 |
| Positive end-expiratory pressure ($cmH_2O$) | 5 ± 0 |
| End-tidal carbon dioxide (mmHg) | 36.9 ± 0.9 |
| Peak inspiratory pressure ($cmH_2O$) | 17.8 ± 1.3 |
| Plateau pressure ($cmH_2O$) | 16 ± 1.4 |

IBW, ideal body weight.

**Supplemental Table 3. Correlation of mean RBC velocity with density and flow microcirculatory variables**

| | RBC velocity | Consensus PPV (small) | Consensus PPV | De Backer score (small) | De Backer score |
|---|---|---|---|---|---|
| RBC velocity | | **0.07** | -0.02 | **0.26** | **0.2** |
| Consensus PPV (small) | **0.07** | | **0.84** | **0.19** | **0.32** |
| Consensus PPV | -0.02 | **0.84** | | -0.13 | **0.04** |
| De Backer score (small) | **0.26** | **0.19** | -0.13 | | **0.68** |
| De Backer score | **0.2** | **0.32** | **0.04** | **0.68** | |

Red color: statistically significant positive correlations; Black bold: positive correlations.
